# Augmenting Structured Diagnoses through Effective Use of Pre-trained Large Language Models on Clinical Notes

**DOI:** 10.64898/2026.05.30.26354533

**Authors:** Hanieh Razzaghi, Nhat Nguyen, Mohan Pargi, Kaleigh Wieand, H. Timothy Bunnell, L. Charles Bailey

**Author notes:** **Corresponding author:** Hanieh Razzaghi, PhD, MPH, Roberts Center for Pediatric Research, 2716 South St, Philadelphia, PA 19146.

## Abstract

**Objective:** Clinical narrative provides a unique window into provider reasoning and attribution, but use has been limited by resource requirements and extensive fine-tuning, and LLMs in particular have traditionally not performed well at medical coding. We optimize and evaluate a reproducible method for automated diagnosis assignment using LLMs in clinical notes and compare with EHR structured diagnoses.

**Methods:** We used GPT-OSS for prompt engineering and task segmentation to create a model that extracts ICD-10-CM diagnoses, with estimates of severity, currency, and importance, from progress notes. We assessed performance across multiple cohorts of patients aged 0-21 years. For each, 100 outpatient provider notes were selected across levels of severity, along with coded diagnoses from that visit (EHR); a subset of 130 notes were subjected to clinical expert review.

**Results:** Comparison showed 18.7% exact code and 33.3% ICD-10-CM category match between EHR and LLM, but semantic similarity of 0.93 at the category level. Compared to expert review, LLM precision was 0.84 and recall 0.49 for exact matches, and 0.92 and 0.62, respectively, for category-level matching. In contrast, EHR coded diagnoses showed slightly higher precision (0.94 for both cases) and substantially lower recall (0.27 and 0.43) versus expert review. Codes not identified by the LLM were more often rated by the reviewer as lower importance or certainty.

**Conclusion:** We demonstrate a reusable approach to optimizing a pretrained LLM for use in diagnosis extraction from clinical notes, facilitating large-scale diagnosis screening by LLMs without the need for expensive study-specific model refinement.

## BACKGROUND AND SIGNIFICANCE

Secondary use of clinical data offers an unprecedented opportunity to conduct research at scale[1-6]. Multi-institutional networks have accelerated research using Electronic Health Record (EHR) data, but most of these networks rely heavily on structured data to derive patient insights and study health outcomes[7-10]. However, the use of unstructured data for augmenting EHR structured data has become increasingly widespread and shows promise to transform clinical research at scale[11,12] . Pediatric research may uniquely benefit from routine use of unstructured data, as measures of well-being and functioning may not be adequately captured in the structured data alone. Provider clinical narratives in particular offer the potential to capture more comprehensive data about patient medical histories[13,14]. Significantly, patient diagnoses and other clinical events can be annotated by provider notes to contextualize scope, severity, and significance of a patient’s medical status. These metadata provide a more complete understanding of patient disease status than recorded diagnoses alone[15]. For example, asthma is a common diagnosis in children, but severity and exacerbation are difficult to capture across time, with structured diagnoses offering only a limited window into patient clinical histories. Proxy measures such as diagnosis frequency and medication use can approximate disease status, but these measures are often incomplete and based on unverified assumptions (e.g., multiple instances are related to severity). This is further complicated by billing requirements motivating choice of structured codes, which can obscure clinical intent or motivation during an encounter.

Analysis of clinical text has traditionally relied on NLP methods that are time-intensive to deploy, require substantial effort for note segmentation, and have limited capacity to complete complex tasks beyond assertion and negation[16-18]. Further, direct note-to-code tasks remain a significant challenge[19]. Large Language Models (LLMs) offer the potential to enhance clinical research by ingesting large volumes of unstructured clinical text to support tasks such as evidence summarization and synthesis[20,21]. LLMs are already increasingly deployed within health systems to leverage this capacity through discharge summary generation and ambient documentation, which reduces burden on clinicians[22,23]. Drawing from these strengths, LLMs have also been applied to clinical notes for research to extract clinical concepts without extensive task-specific training, enhancing and complementing data derived from EHR-structured data[24]. However, this research has been limited to symptom identification and summarization in domain-specific settings or a small or constrained set of tasks[25-27]. Scaling this to more complex tasks such as LLM assignment of diagnostic codes has historically resulted in poor performance and conclusions that LLMs demonstrate substantial performance gaps[28,29]. Fine-tuning may improve performance, but is also time-intensive and risks over-fitting[30,31]. Further, there are no studies to our knowledge evaluating how LLM code-assignment may augment or compare with structured EHR data.

## OBJECTIVES

We seek to address this critical gap by assessing LLM performance in diagnostic code assignment from clinical provider notes. We highlight the value of notes by comparing LLM output to clinician-assigned structured codes, demonstrating that clinical notes augment information derived from structured EHR data alone. Finally, we validate both LLM diagnoses as well as the extracted structured diagnosis codes with expert review.

## METHODS

### Population

Data was drawn from a corpus of clinical notes available to the study team at Children’s Hospital of Philadelphia (CHOP), described in detail elsewhere[32] (IRB 21-019569). These notes were extracted from patient records who had evidence of testing or immunization for SARS-CoV-2 infection, COVID-19 diagnosis code, or other respiratory illness between January 2019 and December 2022. Our study sample was selected to comprise patients with varying degrees of medical complexity to better assess the capacity of LLMs to accurately recognize diagnoses across different note structures. Therefore, we identified patients with COVID-19, inflammatory bowel disease (IBD), or asthma, insuring representation of patients with complex chronic conditions according to the pediatric medical complexity algorithm (PMCA)[33].

We required the following cohort-specific criteria: (1) Asthma: two asthma diagnoses 90 days apart with one bronchodilator prescription (n=100 patients); (2) IBD: three or more Crohn’s disease diagnosis and one medication based on the phenotype previously developed in PEDSnet[34] (n = 100 patients) as well as one IBD-related lab; (3) COVID-19: positive RT-PCR or antigen test for SARS-CoV-2 and symptoms associated with COVID-19 at the same visit (n = 100 patients). For the first two categories, we additionally required a diagnosis classified as a progressive disease according to the PMCA for 50 patients from each cohort. Additionally, for the COVID-19 cohort, we required that 50 patients were hospitalized within 7 days of COVID diagnosis & on mechanical ventilation or prescribed a steroid, remdesivir, or nirmatrelvir/ritonavir during hospitalization. We also imposed the following criteria to be included in the final analyses: (1) At least five encounters between 2015 and 2024 at CHOP, with minimum two outpatient encounters; (2) First and last clinical note should be at least two years apart; (3) Structured data includes at least: one diagnosis outside of the inclusion chronic condition (i.e., COVID-19, IBD, Asthma), one procedure, and one drug prescription. We limited all analyses to outpatient progress or H&P note types with at least 2000 characters.

### LLM Selection and Computing Environment

We performed initial pilot testing on the open-source version of GPT (OSS) 120B model[35]. The model was deployed using Ollama[36], an open-source LLM inference engine, on dual NVIDIA A100 80GB GPUs. VRAM utilization ranged from 80GB to 160GB depending on batch size and sequence length. To accommodate variable unstructured note lengths encountered

in clinical processing pipelines, the context window parameter was dynamically scaled to the next power-of-2 context window size: 4096 tokens for notes with lengths between 1024 and 2048 tokens, and 8192 tokens for notes between 2048 and 4096 tokens. This KV cache-optimized strategy ensures complete capture of clinical content without truncation, providing sufficient headroom for zero-shot diagnosis extraction, chain-of-thought reasoning, and importance scoring across the full spectrum of clinical note requirements. We set the temperature parameter to be 0 for the LLM to generate consistent results.

We initially evaluated responses to prompts instructing the models to agree or disagree with EHR clinician-assigned diagnoses. We reviewed model results based on fidelity to prompt instructions (e.g., formatting output) as well as correctness (e.g., final decision more accurate).

### Prompt Engineering

Prompts were developed using a zero-shot approach with guidance to align model outputs with the desired output structure. The initial prompt set emphasized clear but relatively undirected instructions. We first piloted these prompts on a subset of notes to refine the framing and improve response quality. A split prompting strategy was then adopted, where the initial prompt targeted diagnosis code extraction and subsequent prompts focused on metadata extraction from the clinical notes, including code classification (current, history, mentioned), importance (low, medium, high), and severity level (low, medium, high).

The subsequent prompt iteration improved upon initial versions through task-specific, definitive prompting that precisely defined each metadata label to enable accurate LLM categorization of clinical codes. System prompts were tailored to individual tasks, while user prompts incorporated both definitions and input data, following the multi-stage process outlined in Figure 1. First, an LLM extracted diagnosis codes from the clinical note; subsequently, three independent LLMs received these codes along with the original note to classify each code (Reason for Visit, Other Active, Possible, History, or Mentioned conditions), assess importance (prominence of extracted diagnosis relative to the note), and evaluate severity/patient impact (defined as potential for adverse outcomes). Metadata categories were refined after initial expert review to more accurately capture characteristics of diagnoses. Responses were constrained to structured JSON format with keys for classification, importance, severity, and rationale. The complete prompts appear in Table S1.

**Figure 1.**
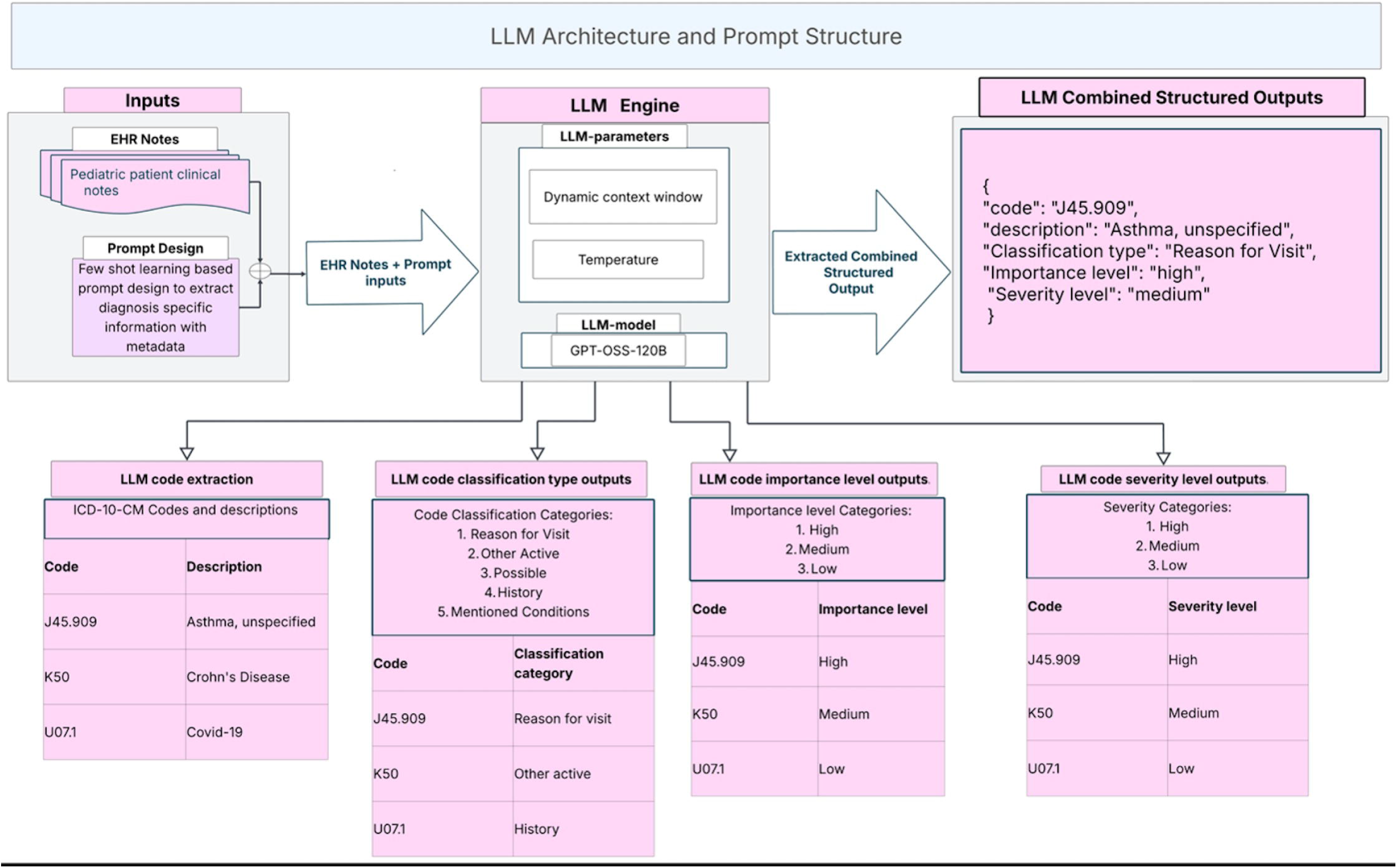
LLM Architecture. Flow diagram illustrating multi-stage LLM & prompting architecture.

### Clinician Reviews

We conducted clinical expert reviews to validate the precision and recall of both the LLM-derived diagnoses (LLMdx) extracted from clinical notes as well as the structured clinician-assigned diagnoses (EHRdx) associated with the visit. The reviewer was a board-certified pediatrician and informatician (LCB). The review sample comprised 130 de-identified clinical notes from distinct patients drawn equally from the three cohorts. For this subset of notes, we required that the note length be ≥ 2000 characters and the number of EHRdx be between one and seven unique diagnoses. To augment the possible diagnosis set for each note, and thus reduce risk of an inflated recall statistic, we prompted a separate LLM to extract potential diagnoses from the notes by referencing a Retrieval-Augmented-Generation (RAG) data store containing ICD-10-CM diagnosis codes and terms. We also tested tools such as medspaCy for feature extraction, but the LLM and RAG store combination (RAGdx) provided more comprehensive and relevant ICD-10-CM codes during testing.

We developed a REDCap case report form in which the reviewer assessed the accuracy of diagnosis codes and independently assigned values in the same metadata categories as the LLM. For each note, the reviewer was presented with the deduplicated set of LLMdx, EHRdx, and RAGdx, and was blinded to the source. The reviewer was prompted to verify whether the diagnosis could be derived from the note, and to classify the code as current diagnosis, part of the patient’s history, or just mentioned in passing. If the diagnosis was determined to be current, the reviewer was then asked whether it was the primary reason for the visit. For each confirmed diagnosis, the reviewer was prompted to assign importance (high, medium, low) and severity scores (high, medium, low). Finally, the reviewer added to the form diagnoses that they judged were indicated in the note but were missing from the combined list.

### Comparison of LLMdx and EHRdx

All diagnoses were represented as ICD-10-CM codes. We performed comparison across sets of codes using exact code matching, category matching (matching of all characters up to the ‘.’ in each ICD-10-CM code), and chapter matching (matching of the leading alphabetic character).

We computed the median (IQR) and mean (SD) number of diagnoses per note for each method, as well as distributions of the metadata terms for LLMdx (i.e., reason for visit, importance, severity). To compare the overlap of LLMdx and EHRdx for the full cohort, we compared the intersection of LLMdx and EHRdx by exact code and ICD-10-CM category match. We also computed semantic similarity scores using two established similarity methods from the *simona* R package[37]: (1) *Wu-Palmer[38]*, which calculates relatedness of two terms by considering the depths in a hierarchical taxonomy and their nearest common ancestor, and, (2) *Leacock-Chodorow[39]*, which calculates relatedness based on shortest path between them, rather than to the nearest ancestor. To calculate an overall similarity score for each encounter, we computed the Best Match Average (BMA), which finds the highest similarity score for each diagnosis code from each source and computes the average[40]. Specifically, we implemented the formula below:

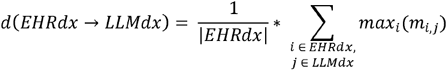

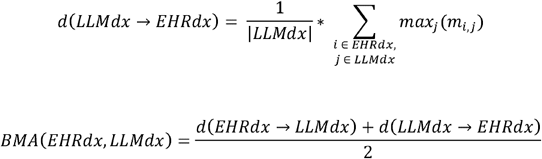

where :

d(EHRdx→LLMdx) = directional best match every point in set EHRdx to the nearest point in set LLMdx

d(LLMdx→EHRdx) = directional best match every point in set LLMdx to the nearest point in set EHRdx

m_*i,j*_ = semantic similarity (either Wu-Palmer or Leacock-Chodrow) between EHRdx□ and LLMdx□

BMA scores were computed using complete ICD-10-CM codes as well as codes truncated at the decimal point, yielding similarity between subtrees representing variations on a particular diagnosis.

### Validation of LLMdx and EHRdx

To independently assess the accuracy of LLMdx and EHRdx with respect to each specific note, we compared against the clinican review gold standard (CRdx). Specifically, we computed recall and precision both for exact code match as well as matches that overlap based on ICD chapter code, using the following formulas, replacing LLMdx with EHRdx appropriately:

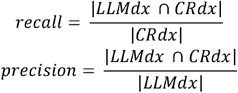

In addition to the overall performance, we computed agreement of metadata related to importance and relatedness to visit. We report both overall and sub-analyses in the results.

### Data Availability

The results reported in this manuscript are based on detailed individual-level patient data. Due to the high risk of reidentification based on unique patterns in the clinical data, even when demographic identifiers have been removed, patient privacy regulations prohibit us from releasing the individual-level data publicly. Requests to reuse the data, with appropriate research ethics and data sharing arrangements, may be made to PEDSnet. Please direct requests to access the data, either for reproduction of the work reported here or for other purposes, to.

### Code Availability

The code used in this study is available on GitHub and can be accessed at https://github.com/PEDSnet/llm_dx_note_extract

## RESULTS

We present characteristics of the full and clinician reviewed cohort in Table 1. The clinician review cohort consists of notes from distinct patients, while the notes cohort includes multiple notes per patient in the cases where there were insufficient patients with the required eligibility condition and severity of comorbid conditions. Consequently, the disease-specific sub-cohorts are represented equally in each of the clinician reviewed and notes cohort. Black patients were over-represented in the clinician reviewed cohort, but other demographic characteristics were similar.

**Table 1.** Cohort Characteristics.

|  | Clinician review cohort<br>(N=130) | Notes cohort<br>(N=243) |
| --- | --- | --- |
| Age [Q1, Q3] | 8 [4.2, 13.1] | 6.2 [3.8, 14.2] |
| Sex |  |  |
| Female | 67 (51.5%) | 117 (48.1%) |
| Male | 63 (48.5%) | 126 (51.9%) |
| Unknown | 0 (0%) | 0 (0%) |
| Race |  |  |
| American Indian/Alaska Native | 3 (2.3%) | 1 (1.3%) |
| Asian/Native Hawaiian/Other Pacific Islander | 25 (19.2%) | 10 (12.5%) |
| Black | 38 (29.2%) | 17 (21.3%) |
| Multiple Race/Other/Unknown | 24 (18.5%) | 12 (15.0%) |
| White | 40 (30.8%) | 40 (50.0%) |
| Ethnicity |  |  |
| Hispanic | 13 (10.0%) | 20 (8.2%) |
| Not Hispanic | 117 (90.0%) | 221 (90.9%) |
| Unknown | 0 (0%) | 2 (0.8%) |
| Number of notes per patient |  |  |
| 50 | 22 (16.9%) | 36 (14.8%) |
| 51-100 | 32 (24.6%) | 47 (19.3%) |
| 101-200 | 32 (24.6%) | 63 (25.9%) |
| 201+ | 44 (33.8%) | 97 (39.9%) |
| Disease-specific cohort |  |  |
| IBD | 43 (33.1%) | 80 (32.9%) |
| Asthma | 43 (33.1%) | 82 (33.7%) |
| COVID-19 | 44 (33.8%) | 81 (33.3%) |

### 3.1 Refinement of LLM Feature Extraction

The diagnosis codes and metadata extracted from the LLMs were reviewed by the study team to adjudicate discordant cases. We first observed that when we used only a zero-shot with a single prompt, diagnosis-related information was not reliably captured. This was likely because the volume of information in a single evaluation was greater than the model could reliably extract. We then observed that more context and specificity were required to label diagnoses to accurately assign what was present for a patient. For example, one snippet from a clinical note in the sample stated: “Left click noted, ultrasound ordered to evaluate for developmental dysplasia of the hip.” The LLM assigned the ICD-10-CM code as Q65.0 (Congenital dislocation of hip), but a clinician expert disagreed, noting that the condition was being evaluated but not assigned to the patient. This discordance highlighted the need for an additional category to capture diagnoses that are possible but not yet established. Consequently, we refined our classification schema to include more granular categories, yielding the final structure described in the methods. In this example, the added category of “possible” diagnosis allowed for semantic alignment. We also found that use of split prompting, that is, serial evaluation for each metadata category by a different model, achieved greater concordance with clinician review than use of a single combined prompt.

### LLM and EHR Agreement

The mean (SD) number of diagnoses per note in the full set of notes for EHRdx, LLMdx, and RAGdx are 3.0 (±1.2), 3.7 (±4.6), and 5.1 (±6.0). The R (signs, symptoms, and laboratory findings) and Z (healthcare utilization) codes had the highest proportion of codes identified by both EHRdx and LLMdx, followed by J (respiratory), K (digestive), and Q (congenital abnormalities) codes, consistent with our cohort definitions.

The exact and category match agreement between these overall sets of diagnoses was 18.7% and 33.3%, respectively. Figure 2 below shows the overall agreement and kappa scores between the LLM-extracted diagnoses and those assigned by the clinician at the visit. We reviewed discrepancies between EHRdx and LLMdx where the kappa scores were lowest. Within the M chapter, the code category most often identified by LLMdx but missed by EHRdx was M41 (Scoliosis). Conversely, EHRdx did not have a concentration of codes missed by LLMdx, but rather, included a range of three category codes missed by LLMdx at low frequency (n=1) and reflect less specific diagnoses: M43 (Other deforming dorsopathies), M79 (Other and unspecified soft tissue disorders, not elsewhere classified), and M85 (Other disorders of bone density and structure). Within the Z chapter, Z93 codes (Artificial opening status) were often identified by LLMdx but not EHRdx, which encompasses tracheostomies and colostomies. In contrast, Z20 (Contact with (suspected) exposure to communicable diseases) were noted by clinicians but not extracted from the text by LLMs. Within the R chapter, LLMdx identified a relatively large number of codes classified under R62 (Lack of expected normal physiological development in childhood and adults). Similar to the M chapter, EHRdx classifications missed by LLMdx were dispersed, but the most frequent was R63 (Symptoms and signs concerning food and fluid intake).

**Figure 2.**
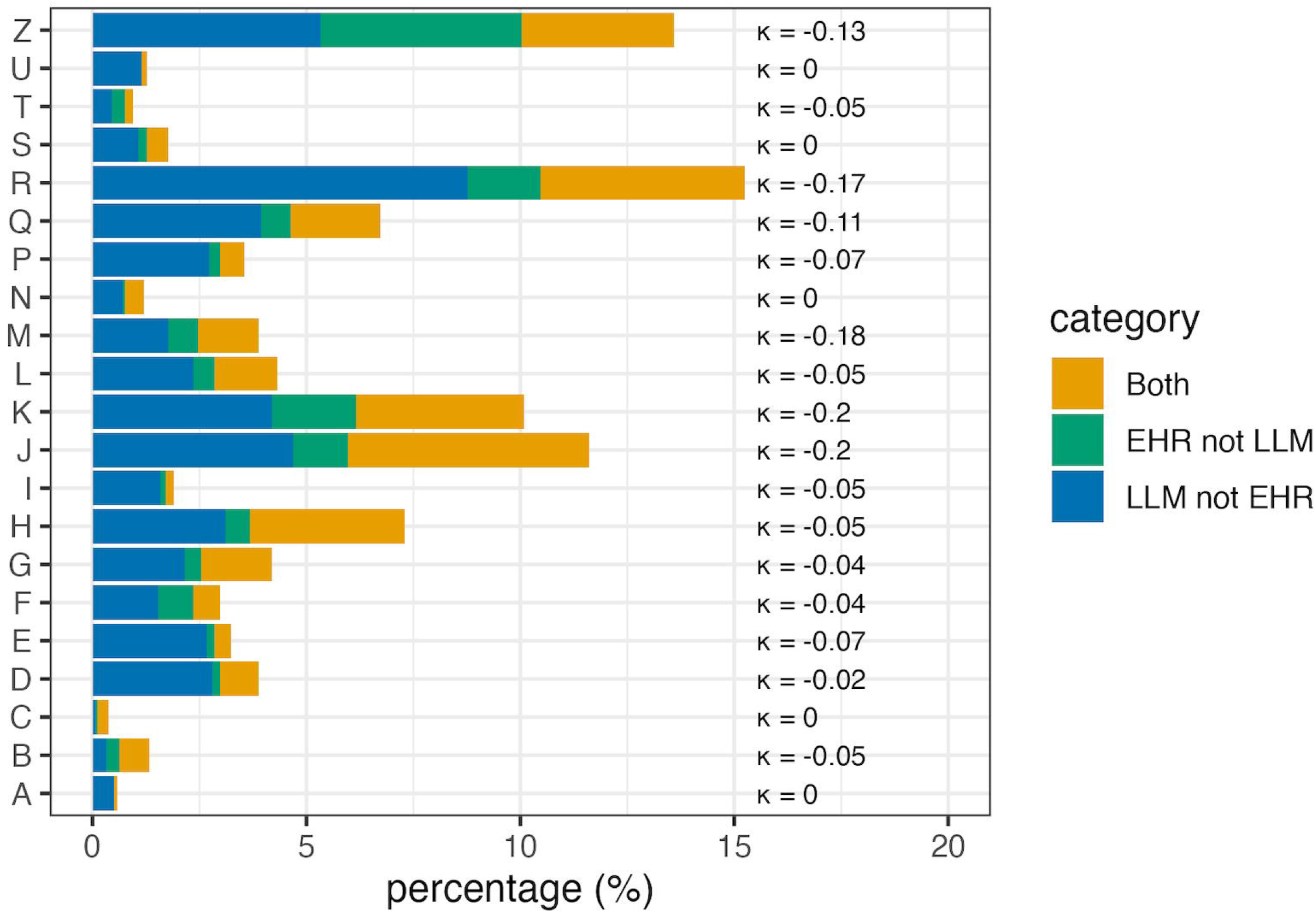
Code Overlap by ICD-10-CM Chapter. Overlap of LLM-assigned and clinician-coded diagnosis assignment for the notes set by chapter match.

The semantic similarity output is shown in Table 2 for all diagnoses, then restricted to those that the LLM assigned as “Reason for Visit” and “High/Medium Importance”. As expected, agreement was higher for ICD-10-CM category and for codes designated as relevant to the current visit or of higher importance. Importantly, the semantic similarity scores demonstrate that while there are notable code-level differences between LLMdx and EHRdx, the discrepancies may most often be due to coding preference for similar concepts rather than significant clinical differences.

**Table 2.** Best-Match Average (BMA) semantic similarity scores using two different similarity scores metrics for notes set (n = 300)

|  |  | BMA Median (95% CI*) |  |  |
| --- | --- | --- | --- | --- |
| Semantic Similarity Method |  | Overall | Reason for visit | High/medium importance |
| Wu-Palmer | Exact | 0.66<br>(0.58 – 0.71) | 0.73<br>(0.67 – 0.75) | 0.75<br>(0.71 – 0.77) |
| Leacock-Chodorow |  | 0.59<br>(0.57 – 0.65) | 0.68<br>(0.64 – 0.73) | 0.77<br>(0.72 – 0.79) |
| Wu-Palmer | Category | 0.93<br>(0.90 – 0.94) | 0.94<br>(0.92 – 0.96) | 0.99<br>(0.96 – 1.00) |
| Leacock-Chodorow |  | 0.89<br>(0.86 – 0.90) | 0.90<br>(0.89 – 0.93) | 0.99<br>(0.93 – 1.00) |

\* 95% CI computed by bootstrapping for 1,000 iterations

### Clinician Review Validation

Table 3 shows the performance of LLM-assigned or clinician-coded diagnoses compared with the clinician review. The EHRdx codes demonstrated high precision, outperforming LLMdx for exact code classifications. However, precision for LLMdx improves for code categories, either comparable with or exceeding EHRdx overall and across different stratifications. Precision was highest for LLMdx when restricted to just codes that were classified as “Reason for Visit” without observing an associated drop in recall. The LLMdx significantly outperformed EHRdx in recall for both exact and category codes, trending higher for the latter, though it never exceeds 67.01%. While LLMdx performs best when restricted to “Reason for Visit,” the only notable increase from its overall performance is for exact code precision.

**Table 3.** Performance Statistics of EHRdx and LLMdx when compared with gold standard.

|  |  | Exact Code | Category |
| --- | --- | --- | --- |

| Scenario | Model | Precision | Recall | Precision | Recall |
| --- | --- | --- | --- | --- | --- |
| **All Codes** | EHR | 93.79 | 27.06 | 94.22 | 43.29 |
| **All Codes** | LLM | 84.01 | 49.23 | 92.24 | 61.83 |
| **LLM & CR: Reason for visit + Other active + History** | LLM | 85.1 | 49.67 | 92.86 | 62.31 |
| **LLM & CR: Reason for visit + Other active** | LLM | 86.64 | 50.4 | 94.07 | 64.97 |
| **LLM & CR: Reason for visit** | LLM | 91.78 | 49.69 | 96.43 | 64.54 |
| **LLM & CR: History** | LLM | 78.23 | 48.45 | 87.7 | 59.45 |
| **LLM & CR: Medium & High Importance** | LLM | 87.33 | 49.43 | 95.35 | 64.13 |
| **LLM & CR: Medium & High Severity** | LLM | 85.32 | 48.98 | 92.03 | 63.29 |
| **LLM & CR: Medium & High Severity + Reason for visit** | LLM | 90.57 | 51.11 | 95.05 | 67.01 |
| **LLM & CR: Medium & High Importance + Reason for visit** | LLM | 91.67 | 50.23 | 96.38 | 65.93 |
| **LLM & CR: Medium & High Severity + Medium & High Importance + Reason for visit** | LLM | 90.57 | 50.84 | 95.05 | 66.67 |

We reviewed diagnosis codes that were identified by RAGdx but not by LLMdx as shown in Figure 3 to understand where the latter was omitting diagnoses. Most codes that were omitted and also verified by the reviewer were classified by RAGdx as “Other Active” or “History” codes, and the vast majority were either “low” or “medium” importance.

**Figure 3.**
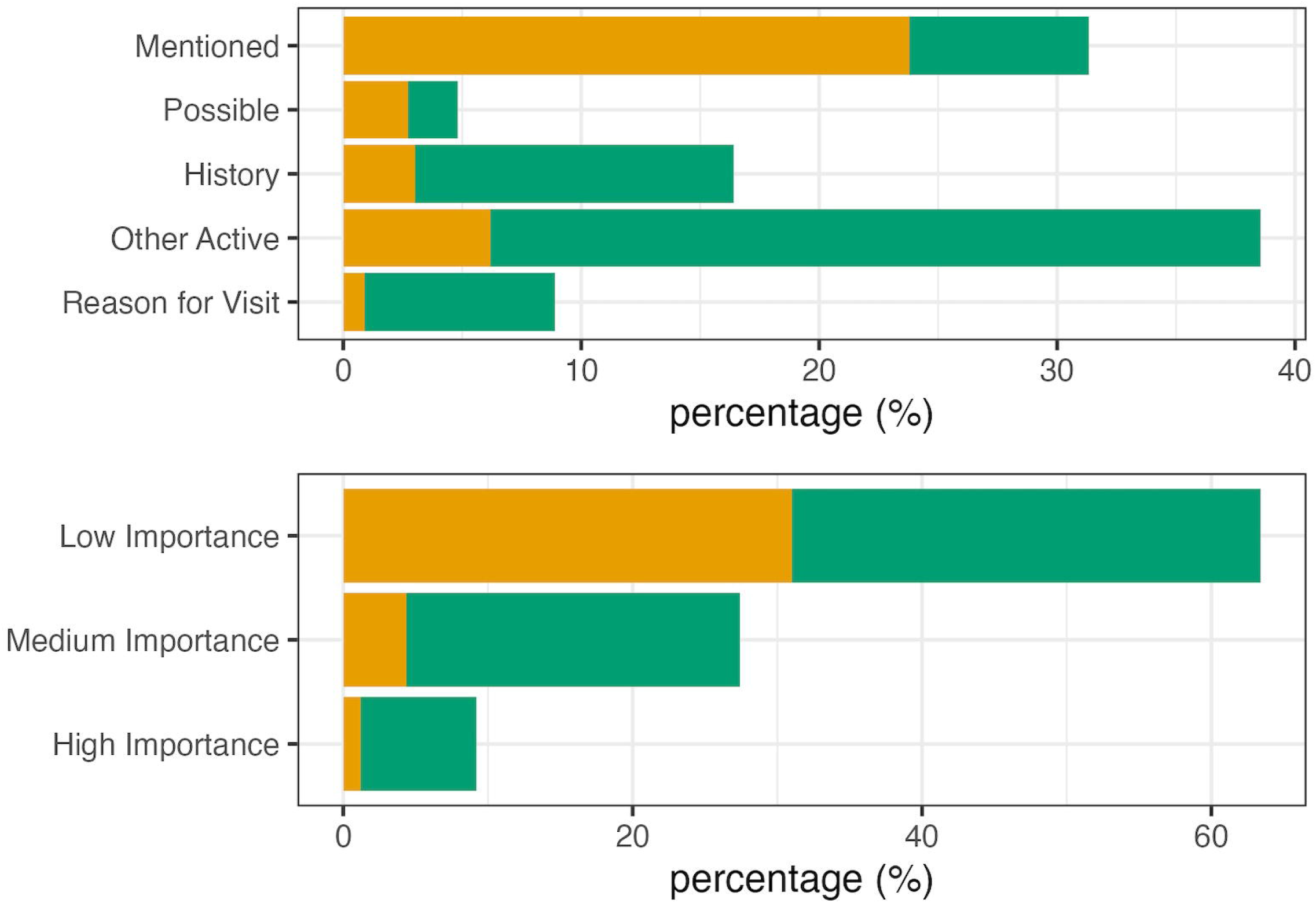
Discordant Metadata Distributions. Metadata distributions for codes that were omitted by LLMdx and captured by RAGdx where green bars represent reviewer agreement and orange bars represent reviewer disagreement

Figure 4 shows the distribution of codes by chapter identified by the RAGdx and verified by the reviewer but missed by LLMdx. The RAGdx was more likely to capture signs and symptoms than clinical diagnoses relevant to patient care. The most common R codes captured only by RAGdx include R52.9 (Pain, unspecified), R50.9 (Fever, unspecified), and R06.2 (Wheezing). Taken together, results indicate that LLMdx performed well for clinical codes most relevant to the visit but missed mentions of symptoms also included in the documentation.

**Figure 4.**
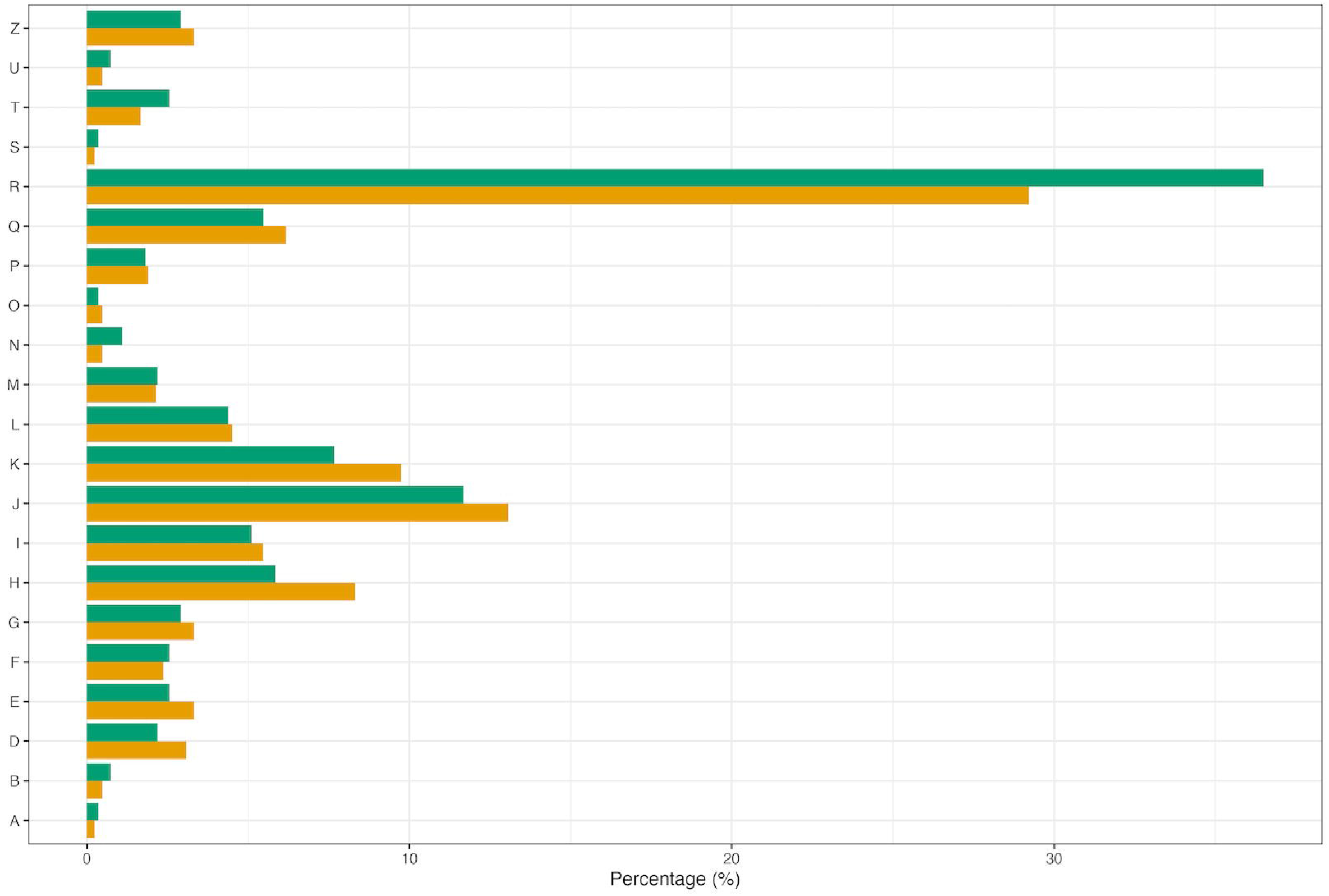
Discordant Code Distribution by ICD-10-CM. Chapter Distribution of codes omitted by LLMdx and captured by RAGdx that reviewer agreed to for exact and prefix matching, where green bars represent concordance for code categories and orange bars for exact code.

## DISCUSSION

In this study, we evaluated the ability of LLMs to extract diagnosis codes directly from clinical notes, comparing LLM-derived diagnoses (LLMdx) against both structured EHR diagnoses (EHRdx) and clinician review. Our findings demonstrate that LLMs can achieve high diagnostic code extraction performance without task-specific fine-tuning, but instead through prompting strategy and model architecture. Although LLMs and clinicians assigning visit diagnoses tended to select different codes, alignment on clinical entities was strong.

LLMdx demonstrated generally high performance in assigning diagnosis codes directly from unstructured clinical notes, suggesting that LLMs possess sufficient clinical language encoding to support diagnostic code extraction in a zero-shot or few-shot setting. However, our findings underscore that prompting strategy must be highly specific and directive. This is consistent with the broader literature on prompt engineering, which has demonstrated that LLMs are sensitive to the framing, specificity, and structure of instructions provided[41,42]. However, our study provided prompting and architecture strategies that outperformed previous attempts to generate coding assignments through LLMs[28,43].

A particularly important finding was the role of metadata terms in optimizing LLMdx performance. For each extracted code, we assigned metadata along three dimensions: purpose (reason for visit, mentioned, possible, history, other active), importance (high, medium, low), and severity (high, medium, low). We improved prompts through metadata contextualization and providing highly specific tasks, which guided the model toward more accurate and clinically relevant code assignment. This represents a meaningful opportunity over EHRdx, which is generated through structured clinical workflows and cannot be easily interpreted post hoc. The ability to modulate LLMdx output through metadata offers investigators a degree of analytical flexibility that structured EHR data does not readily afford.

Agreement between LLMdx and EHRdx was better when codes were measured at the category level rather than exact code matches. This level of specificity is often sufficient for research variables, which are less likely to depend on information such as anatomic location conveyed by the portion of ICD-10-CM codes after the point. Moreover, semantic similarity metrics demonstrated marked improvement in overlap. Discrepancies between LLMdx and EHRdx may also reflect differences in the purpose and process of code assignment between the two approaches. EHRdx is generated within a billing and administrative workflow, and as such tends to capture administrative and procedural or chronic condition codes that may not reflect the full clinical narrative of an encounter. LLMdx, on the other hand, was guided by prompts explicitly focused on clinical diagnoses and tended to prioritize clinically significant entities documented in the note. This is demonstrated in the EHRdx selection of Z-codes that focused on billing requirements, which LLMdx often omitted. By contrast, EHRdx omitted diagnostic codes for more descriptive clinical diagnoses.

When evaluated against the clinician review, EHRdx demonstrated generally high precision for exact and category code assignment but substantially limited recall, consistent with the known incompleteness of structured administrative data where many clinically relevant diagnoses documented in the notes are never formally coded^40,41^.

LLMdx across all codes had better recall for exact code assignment with a modest reduction in precision. This pattern was even more pronounced at the category level, where LLMdx achieved a recall of 61.83% compared to 43.29% for EHRdx, with comparable precision. When investigating omissions in recall, LLMdx most often did not contain low importance diagnoses that were related to symptom-based codes.

The utility of the metadata assignments was reflected in meaningful variation in precision and recall across subsets. Restricting LLMdx to the reason for visit metadata category produced the most favorable balance of precision and recall. This suggests that when investigators only wish to assess codes relevant to a specific encounter, constraining extraction to reason for visit codes offers a practical strategy for optimizing precision without substantially sacrificing recall relative to other LLMdx subsets. Filtering by medium and high importance codes produced similarly balanced performance, offering investigators an alternative dimension along which to tune extraction depending on the analytic priorities of a given study. Further, restricting to codes relevant to the current visit with medium or high severity produced the highest recall.

Importantly, LLMdx more often omitted certain codes, particularly those representing signs and symptoms. These codes were more likely to be unrelated to the current visit and assessed as of low importance. Additionally, EHRdx had a higher proportion of codes reflecting “other specified” or “unspecified” clinical entities, possibly reflecting mapping from interface terminologies in use at the point of care. Understanding the systematic patterns of LLMdx omissions, particularly when identified by EHRdx, is an important area for future refinement of prompting strategies.

### 1. Conclusion

These findings have several implications for clinical researchers. First, LLMs represent a viable and flexible approach to diagnosis code extraction from unstructured clinical notes, particularly because structured data may be incomplete, decontextualized (*e*.*g*. by auto-inclusion from prior encounters), or administratively biased. Second, the quality of LLM-derived codes is highly dependent on prompting strategy and LLM architecture, and investment in prompt development and validation is essential before deployment in research pipelines. Third, the metadata dimensions introduced here provide investigators with a tractable mechanism for navigating the precision-recall tradeoff in diagnostic code extraction. A study seeking to maximize case identification, such as estimating disease prevalence, may prioritize code capture by broadening the metadata scope, while one focused on reducing misclassification may constrain extraction to reason for visit or high importance codes to optimize precision. Finally, the complementary strengths of LLMdx and EHRdx suggest that a combined approach may offer advantages over either method alone, and represents a promising direction for future work in clinical data science.

## Data Availability

Patient-specific data cannot be made publicly available due to privacy risk. The code used in this study is available on GitHub.

https://github.com/PEDSnet/llm_dx_note_extract

## Author Contributions

HR contributed to the conceptualization, methodology, analysis and interpretation, drafting of the full manuscript, and integration of all edits. NN contributed to methodology, analysis, data curation, figures and tables, and manuscript editing. MP contributed to the LLM design and implementation, figures, and manuscript editing. KW contributed to analysis, figures and tables, and manuscript editing. HTB. contributed to conceptualization and manuscript editing. LCB contributed to conceptualization, methodology, analysis and interpretation, manuscript editing, and funding. All authors read and approved the final manuscript.

## 2. Acknowledgments

The research reported in this publication was conducted using data from the Children’s Hospital of Philadelphia, a member of PEDSnet, A Pediatric Clinical Research Network. PEDSnet has been developed with funding from the Patient-Centered Outcomes Research Institute (PCORI); PEDSnet’s participation in PCORnet is funded through PCORI award RI-CHOP-01-PS10. The authors are grateful for the sharing of information by the patients, families, and institutions in PEDSnet, which makes more effective research possible. The authors also wish to express their gratitude to Miranda Higginbotham, whose contributions to administrative and regulatory management were critical to this work.

